# Reach, implementation fidelity, and safety of bubble continuous positive airway pressure (bCPAP) therapy in children with severe pneumonia in Pakistan

**DOI:** 10.64898/2026.03.19.26348802

**Authors:** Nadir Ijaz, Ameera Shabbir, Perah Bachal, Haania Rizwan, Muhammad Uzair, Noor Ul Ain, Zareen Qasmi, Ibrahim Shakoor, J. Lucian Davis, Fyezah Jehan, Eric D. McCollum, Qalab Abbas

## Abstract

Bubble continuous positive airway pressure (bCPAP) is a low-cost respiratory support device that has demonstrated different outcomes for children with severe pneumonia in different settings. Some differences in outcomes may be attributable to implementation factors (e.g., patient monitoring and feeding practices). We aimed to characterize bCPAP reach, implementation fidelity, and safety outcomes for children with severe pneumonia in Pakistan.

We conducted a prospective cohort study at Aga Khan University Hospital and Abbasi Shaheed Hospital from February through May 2025. We enrolled children 1–59 months who met WHO criteria for severe pneumonia within 24 hours of presentation to the emergency department. Participants were followed daily via chart review, caregiver survey, and physical exam through discharge, transfer, or death. We reported the proportion of children receiving bCPAP (“reach”) and constructed a mixed-effects, multinomial logistic regression model with robust standard errors to report: fidelity (child location in a highly monitored area, continuous monitoring, avoidance of unplanned disruptions to bCPAP, and avoidance of oral feeding); safety (aspiration events and pneumothorax); bCPAP failure (death, respiratory support escalation, or leaving against medical advice); and in-hospital mortality.

Of 165 children with severe pneumonia, 88 (53%) received bCPAP over 141 bCPAP days. The average predicted probabilities (95% CI) of our fidelity measures were: 85% (78-92%) for location in a highly monitored area; 56% (51-60%) for continuous monitoring; 66% (57-75%) for continuous bCPAP without disruptions; 46% (36-55%) for avoidance of oral feeding while on bCPAP. Among children receiving bCPAP, 9 (10%) experienced an aspiration event, 1 (2.2%) experienced a pneumothorax; 19 (22%) experienced bCPAP treatment failure. One child (1.1%) died; 6 (6.8%) required respiratory support escalation; 14 (16%) left against medical advice.

We identified several gaps in bCPAP reach and fidelity. These may be modifiable by individual-and team-targeted strategies to reduce bCPAP-related complications and pneumonia-related child deaths.

## Introduction

Worldwide, pneumonia remains the number one cause of death among children aged 1-59 months, causing an estimated 424,000 child deaths globally in 2023 (1). Most deaths occur in children with severe pneumonia requiring oxygen therapy in low- and middle-income countries (LMICs) such as Pakistan, which has the third highest number of pneumonia-related child deaths (2). Optimizing the implementation of cost-effective therapies for children with severe pneumonia in Pakistan and other LMICs is therefore urgently needed.

Bubble continuous positive airway pressure (bCPAP) is one such low-cost therapy (3). bCPAP provides oxygen and airway pressure to improve blood oxygenation and work of breathing. In settings with an oxygen or medical air source, bCPAP can be constructed locally using a water canister, respiratory tubing, and nasal interface for as little as $3 USD (3–7). It has been shown to reduce mortality in children aged 1-59 months with severe pneumonia and hypoxemia by as much as 75% compared to low-flow oxygen in multiple trials in LMICs (8–10). However, one large trial in Malawi was stopped early due to increased mortality in children receiving bCPAP therapy compared to those receiving low-flow oxygen therapy (11). Compared to the positive trials, this trial was conducted in a lower-resourced district-level hospital where there was no intensive care capacity, fewer nurses, and only fortnightly rounds by a pediatrician. In the trial, study staff inconsistently adhered to the intended nasogastric tube protocol: only 13% of children eligible for nasogastric tube placement to administer feeds and prevent tracheal aspiration of gastric contents had a nasogastric tube successfully placed (11–13).

These and other bCPAP studies suggest that the context in which bCPAP is implemented at least partially influences its reach (the proportion of eligible children who receive bCPAP therapy) and other outcomes (12,13). Indeed, we have previously conducted a realist review to describe how implementation context (i.e., the local circumstances in which bCPAP is implemented) at five different “levels” may influence bCPAP fidelity (whether bCPAP therapy is delivered as intended) to drive bCPAP safety and effectiveness for children with severe pneumonia in LMICs (14) (**Figure 1**). Some of these levels include factors that are potentially modifiable by individual- and team-targeted strategies and directly affect fidelity by causing disruptions in bCPAP therapy, inadequate patient monitoring, and risky feeding practices. Despite this, few published studies measure the fidelity of bCPAP therapy in children with pneumonia (14).

**Fig 1.**
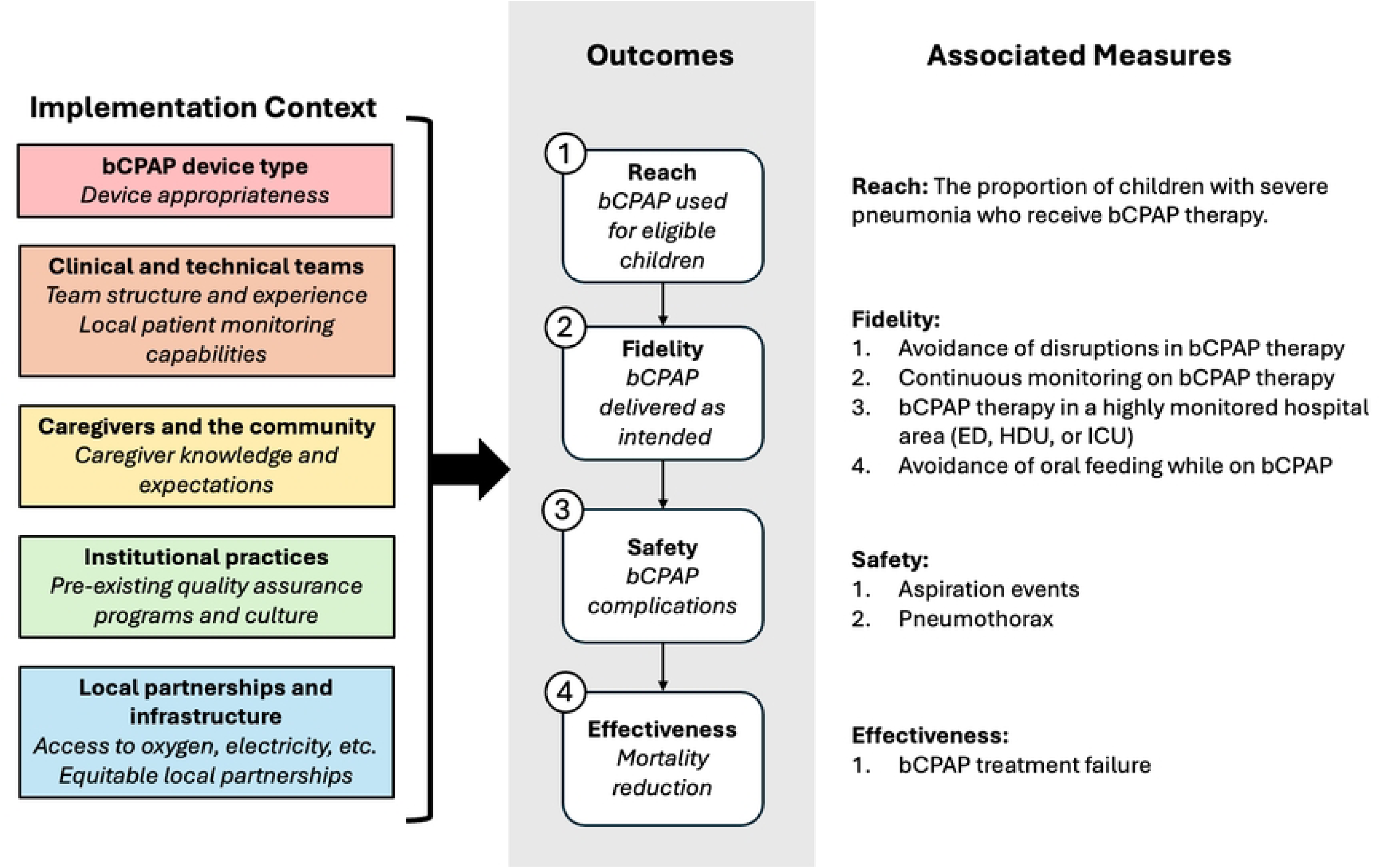
Implementation context and study reach, fidelity, safety, and effectiveness outcome measures. bCPAP = bubble continuous positive airway pressure; ED = emergency department; HDU = high dependency unit; ICU = intensive care unit.

In this prospective cohort study, we aimed to characterize bCPAP reach, implementation fidelity, safety, and effectiveness among children at one private and one public hospital in Karachi, Pakistan.

## Materials and Methods

### Study design & setting

We conducted a multicenter, prospective cohort study at one private and one public hospital in Karachi, Pakistan, intentionally selecting contrasting sites to increase study generalizability. Study sites were Aga Khan University Hospital (AKUH) and Abbasi Shaheed Hospital (ASH). AKUH is a 710-bed tertiary, private teaching hospital. Along with a 22-bed pediatric emergency department, it has 120 inpatient pediatric beds, including 13 pediatric high-dependency unit (HDU) beds and 12 pediatric intensive care unit (PICU) beds. The nurse-to-patient ratio is 1:6 in the pediatric wards, 2:5 in the high-dependency units, and 1:1 in the pediatric intensive care unit. ASH is an 850-bed tertiary, public teaching hospital. Along with a 45-bed pediatric emergency department administered by the ChildLife Foundation (a local non-governmental organization), it has 90 inpatient pediatric beds, including 4 HDU beds and 4 PICU beds. There are two nurses per 40 pediatric ward beds during day shifts and one nurse per 40 ward beds during night shifts. Before this study, both hospitals already used bCPAP locally constructed from a sterile saline bottle and a nasal cannula interface for children with severe pneumonia. In addition, at AKUH, continuous pulse oximetry, high-flow nasal cannula oxygen, non-invasive mechanical ventilation, and invasive mechanical ventilation were readily available. At ASH, additional non-invasive ventilation options were limited, with only 4 mechanical ventilators for children.

Participants were enrolled at AKUH from February 9, 2025, through April 30, 2025, and at ASH from April 7, 2025, through April 30, 2025. Data collection and follow up were completed for all participants between February 9, 2025, and May 5, 2025. This study was approved by the Yale University Institutional Review Board (Protocol Number 2000038591), Ethics Review Committee of the Aga Khan University (Reference Number 2024-10539-31298), the Head of the Department of Pediatrics at Abbasi Shaheed Hospital (ASH), and the Pakistan National Bioethics Committee for Research (Reference Number 4-87/NBCR-1167/24/600). We followed Strengthening the Reporting of Observational Studies in Epidemiology (STROBE) guidelines (15) in reporting our findings (see S1 Appendix).

### Participants

Trained research assistants (RAs) screened for eligibility all children aged 1-59 months who presented to the pediatric emergency department (ED) with a primary chief complaint of cough, difficulty breathing, or respiratory distress. We included children meeting criteria for severe pneumonia, as defined by the World Health Organization (WHO): either cough or difficulty breathing with (1) hypoxemia (SpO2<90%) or central cyanosis, (2) severe respiratory distress, or (3) a general danger sign (vomiting everything, inability to feed, seizure/convulsion, reduced level of consciousness, or deep breathing) (16). To determine study eligibility, children were categorized as having severe respiratory distress if “severe respiratory distress,” “grunting,” or “very severe chest indrawing” was recorded in their medical chart upon ED presentation. Following WHO Integrated Management of Childhood Illness guidelines (17), RAs asked the accompanying caregiver whether their child was vomiting everything, unable to feed, or having convulsions before they presented to the hospital. RAs reviewed the medical chart to assess each child’s level of consciousness on ED presentation.

At AKUH, we included all children who presented to the ED at any time of day if we were able to enroll them within 24 hours of their initial ED presentation. Children at AKUH may have been in the ED or admitted to the ward, HDU, or PICU at the time of enrollment. At ASH, because of higher patient volumes, we included only children who presented to the ED between 9am and 1pm, and these children were all enrolled while they were still in the ED. We excluded children of parents or guardians who did not speak Urdu or English due to a lack of translation and interpretation services for other local languages.

#### Sample size

We targeted a sample size of 165 children. We based this on a preliminary aim to estimate the proportion of children with severe pneumonia who also had hypoxemia (SpO2<90%) in our study population. We assumed a 95% confidence level with a 7% margin of error and an expected proportion of 30% of children with severe pneumonia also having hypoxemia, a conservative estimate based on prior literature (18). While our aims changed during our study due to an inability to collect pulse oximetry data on most children at ASH before oxygen therapy was initiated, we did not change our sample size target, since our remaining aims were descriptive.

## Procedures and data collection

After obtaining written informed consent from the parent or guardian in their preferred language, trained RAs collected participant data using a standardized, secure, web-based survey form in Research Electronic Data Capture (REDCap). All RAs were recent MBBS graduates and received additional training in pediatric physical examination for the purposes of this study. Local certified pediatricians verified and provided feedback for the first five physical exams conducted by each RA before he or she began conducting independent data collection.

On the initial day of enrollment, RAs collected demographic and clinical data via ED chart review, caregiver interview, and physical exam. Demographic data included child characteristics (date of birth and sex) and caregiver characteristics (age, sex, education level, and income). Clinical data included the child’s past medical problems, weight, presenting respiratory symptoms and presence of WHO danger signs, and initial vital signs, as recorded in the child’s chart. RAs measured each child’s mid-upper arm circumference (MUAC). We used the WHO criteria for moderate acute malnutrition (11.5cm ≤ MUAC <12.5cm), severe acute malnutrition (MUAC < 11.5), moderate underweight (−3 ≤ weight-for-age z-score < −2), and severe underweight (weight-for-age z-score < −3) (19).

On both the initial enrollment day and subsequent follow up days, RAs collected additional data on the participant’s current clinical condition (child’s disposition, location in the hospital, and current respiratory support, as applicable) (20). For children who had received bCPAP therapy since ED presentation (on the initial enrollment day) or in the time interval since the last follow-up (on subsequent days), RAs collected additional data regarding bCPAP use. These included bCPAP therapy start times, stop times, and pressure and oxygen concentration settings. The full initial and subsequent day REDCap data collection instruments are available in the S2 Appendix.

Participants were followed up via daily in-person visits until discharge, transfer to another hospital, leaving against medical advice, death, greater than 30 days of admission, or lost to follow-up. At AKUH, participants who could not be physically located in the hospital were deemed lost to follow-up. At ASH, due to high patient volume, RAs requested and used the parent’s or guardian’s contact phone number to follow up participants who could not be physically located in the hospital. For these participants, those without a valid phone number and those who were contacted twice without response were deemed lost to follow up.

## Outcomes

### bCPAP implementation outcomes

We used the RE-AIM framework to guide our study outcome definitions (21). To characterize bCPAP implementation, we focused on bCPAP reach (bCPAP use in our target population, mapping onto the Reach dimension of RE-AIM) and fidelity (bCPAP therapy being delivered as intended, mapping onto the Implementation dimension of RE-AIM). We measured reach as the proportion of children with severe pneumonia who received bCPAP therapy at any point during the study period. We evaluated fidelity by daily assessment of four distinct dichomotous measures. First, we assessed whether continuous bCPAP therapy was delivered without unplanned disruptions (e.g., nasal interface dislodgement, absence of bubbling in water canister, etc.), in the preceding 24 hours by asking bedside nurses and familial caregivers whether bCPAP therapy had been disrupted. While we provided examples of disruptions, any event identified as a disruption by the nurse or caregiver was recorded as such. Second, we assessed whether children on bCPAP were continuously monitored (including at least continuous pulse oximetry, with or without multiparameter cardiorespiratory monitoring). Third, we assessed whether children on bCPAP were in a highly monitored area, defined as an emergency department (ED), high-dependency unit (HDU), or pediatric intensive care unit (PICU). Fourth, we assessed avoidance of oral feeding while on bCPAP by asking bedside familial caregivers whether their child had received any oral feeds over the preceding 24 hours while on bCPAP, regardless of doctor’s orders. We selected these four measures based on our previously published realist review with a diverse panel of 22 bCPAP experts (14), their theoretical importance in maintaining bCPAP therapy effectiveness, and known differences in bCPAP implementation between trials that showed effectiveness and the trial that showed harm.

We collected additional data on reasons for and responses to unplanned disruptions (who recognized and who addressed the disruption) in bCPAP therapy and on feeding practices (diet order, receipt of gastric feeds through a feeding tube, and caregiver administration of additional feeds non-adherent to the diet order).

### Safety outcomes

Our safety and effectiveness outcomes map onto the Effectiveness dimension of RE-AIM. For children receiving bCPAP therapy, we measured: (1) aspiration events, as recorded in the medical chart or reported by the bedside nurse or caregiver after a brief description of how an aspiration event would appear (see S2 Appendix) and (2) pneumothorax, as recorded in the medical chart.

### Effectiveness outcomes

We used a composite bCPAP “treatment failure” outcome, defined as death, escalation of respiratory support to high-flow nasal cannula oxygen or mechanical non-invasive or invasive ventilation, or leaving against medical advice while receiving bCPAP therapy. Our definition of bCPAP treatment failure was based on similar definitions from prior trials (8,10).

## Data analysis

We used descriptive statistics to report child and caregiver demographics for children who did and did not receive bCPAP therapy. We then used descriptive statistics to report bCPAP reach, fidelity, safety, and effectiveness outcomes for the subcohort of children at each site who received bCPAP therapy during the study period. For all variables, we used means and standard deviations to describe normally distributed continuous variables, medians and interquartile ranges to describe other continuous variables, and proportions and frequencies to describe categorical variables. We constructed mixed-effects multinomial logistic regression models with participant-level random intercepts to estimate average predicted probabilities of each fidelity measure over time, accounting for repeated measurements within individuals and adjusting for study site. We used robust standard errors to estimate 95% confidence intervals. Because the proportion of missing data was low (<5% for all variables), we conducted a complete-case analysis. All analyses were conducted using Stata 17 (College Station, TX, USA).

## Results

### Participant enrollment

We screened 177 children, of whom 168 met study eligibility criteria, and 165 whose parent or guardian provided informed consent and were therefore enrolled. Of these 165 children, 157 completed follow up (**Figure 2**). We enrolled 110 children at AKUH and 55 children at ASH. All 8 children who were lost to follow up had presented at ASH.

**Fig 2.**
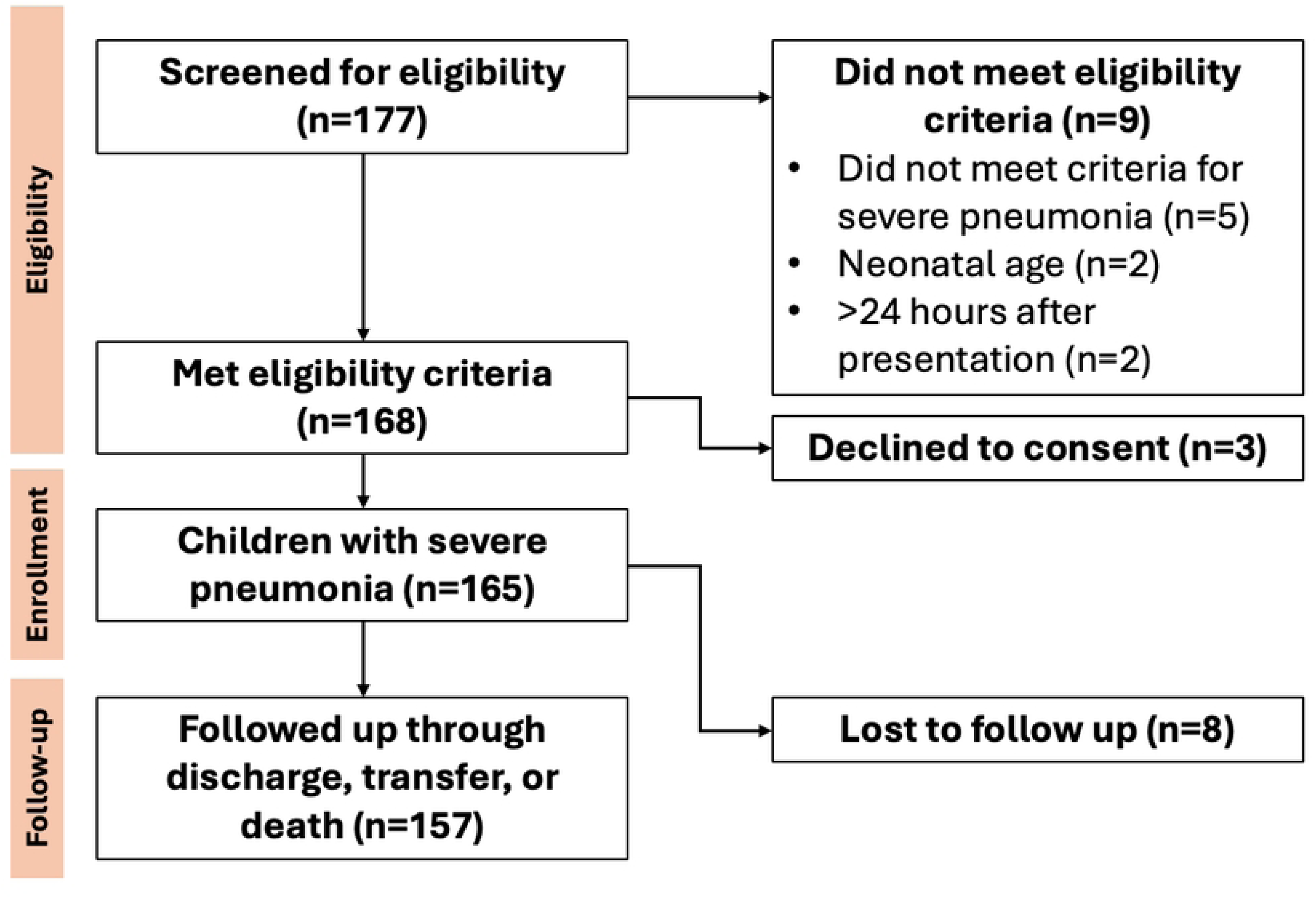
Study flow diagram.

### bCPAP reach and child and caregiver characteristics

At AKUH, 45 (41%) children received bCPAP, 70 (64%) received low-flow oxygen, and 21 (19%) received high-flow nasal cannula oxygen during the study period (patients may have received different types of support at different times during their hospital course). At ASH, 43 (78%) children received bCPAP, 25 (46%) received low-flow oxygen, and 2 (4.4%) received high-flow nasal cannula oxygen during the study period (**Table 1**).

**Table 1.**
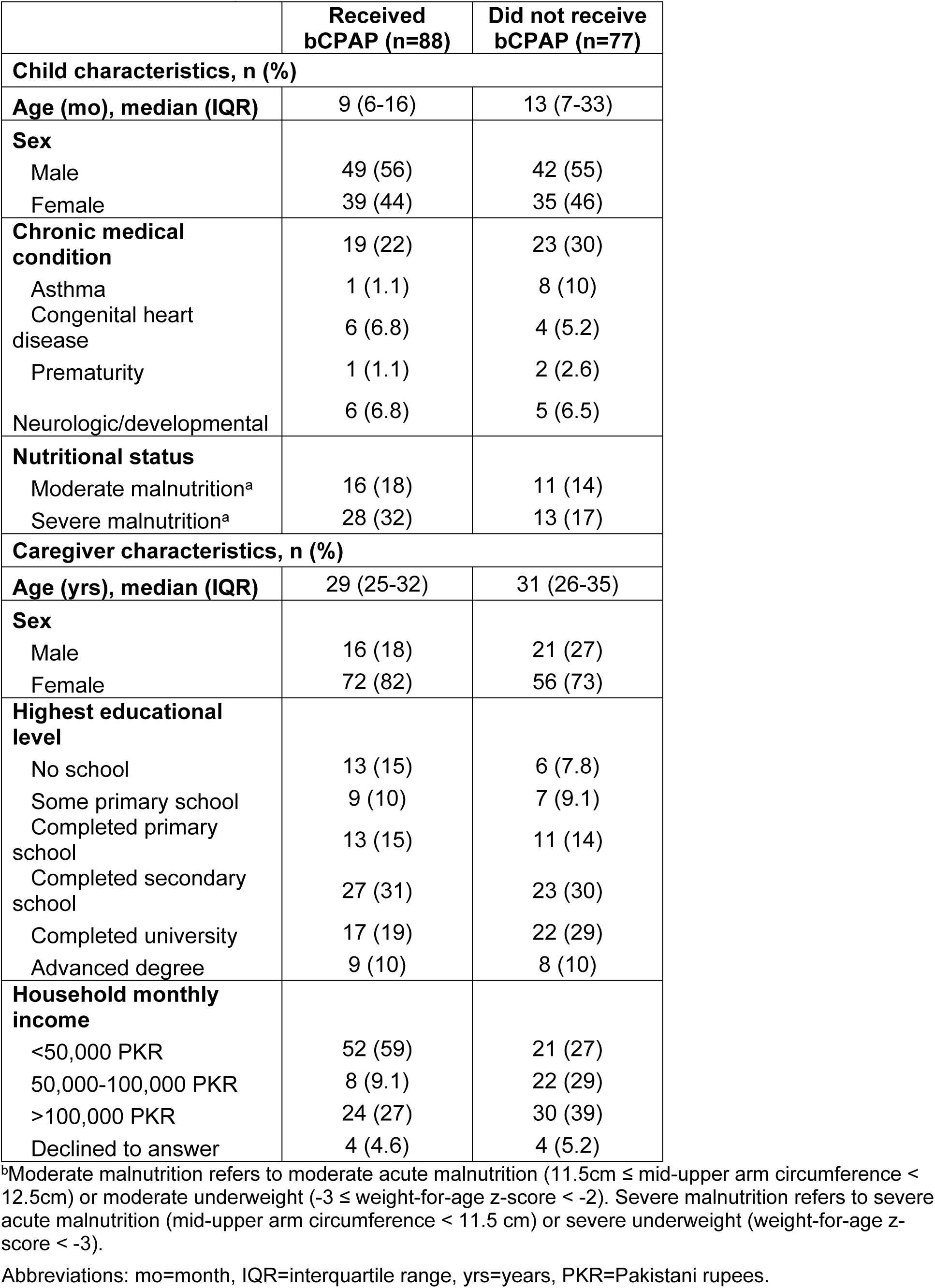
Child and caregiver characteristics for enrolled children who did and did not receive bCPAP therapy.

The median age of children who received bCPAP was 9 months (IQR 6-16 months). Among children who did and did not receive bCPAP, 53 (60%) and 35 (46%) were infants, respectively. Chronic medical conditions were present in 19 (22%) and 23 (30%) children who did and did not receive bCPAP, respectively (**Table 1**). Overall, 44 (50%) children who received bCPAP had moderate or severe acute malnutrition or underweight, with higher rates observed at ASH than at AKUH. Caregivers at AKUH generally had higher educational attainment and household incomes than at ASH. Notably, 45 of 55 (89%) caregivers at ASH reported monthly incomes below 50,000 PKR (approximately $178 USD), compared to 24 of 110 (22%) caregivers at AKUH (see S3 Table for detailed comparisons between sites).

### bCPAP fidelity

Among the 88 children who received bCPAP over 141 bCPAP-days, the average predicted probability of uninterrupted bCPAP therapy was 66% (95% CI 57-75%) (**Table 2**). Among 46 reported unplanned disruptions, the average predicted probability of caregivers identifying the disruption was 80% (95% CI 69-92%); this was 20% (95% CI 9.3-30%) for nurses and 10% (95% CI 1.9-20%) for doctors. The average predicted probability of caregivers resolving the disruption was 54% (95% CI 40-69%); this was 63% (95% CI 50-76%) for nurses and 4.3% (95% CI −1.5-10%) for doctors. Of note, multiple individuals often worked together to identify and resolve disruptions (**Figure 3**).

**Table 2.** bCPAP fidelity outcomes, by study site.

|  | **Average**<br>**predicted**<br>**probability (95%**<br>**CI),<sup>b</sup> AKUH<sup>c</sup>**<br>**(n=75)** | **Average**<br>**predicated**<br>**probability (95%**<br>**CI),<sup>b</sup> ASH<sup>d</sup> (n=66)** | **Average**<br>**predicated**<br>**probability (95%**<br>**CI),<sup>b</sup> Overall**<br>**(n=141)** |
| --- | --- | --- | --- |
| **Undisrupted bCPAP**<br>**therapy<sup>d</sup>** | 67 (57-76) | 66 (50-81) | 66 (57-75) |
| **Monitoring<sup>d</sup>** |  |  |  |
| **Any continuous**<br>**monitoring** | 93 (88-99) | 12 (4.7-20) | 56 (51-60) |
| Pulse oximetry only | 3.2 (-1.2-7.6) | 1.3 (-1.3-3.9) | 2.2 (-0.0026-4.6) |
| Multiparameter monitor | 90 (83-97) | 11 (5-18) | 54 (49-58) |
| **Location** |  |  |  |
| **Any highly monitored**<br>**area** | 86 (76-96) | 84 (75-93) | 85 (78-92) |
| Emergency department | 33 (25-42) | 45 (35-55) | 39 (33-46) |
| High-dependency unit | 49 (39-60) | 39 (26-52) | 44 (36-52) |
| Intensive care unit | 0.028 (-0.014-0.069) | 0.014 (-0.015-0.043) | 0.021 (-0.0038-0.046) |
| **Ward** | 15 (6-24) | 15 (7-23) | 15 (10-20) |
| **Feeding** |  |  |  |
| **Avoidance of oral feeds<sup>d</sup>** | 48 (34-61) | 44 (31-56) | 46 (36-55) |
| **Diet order<sup>d</sup>** |  |  |  |
| NPO | 27 (16-37) | 64 (52-76) | 44 (35-53) |
| Gastric feeds | 25 (15-36) | 6.9 (1.6-12) | 18 (11-25) |
| Oral feeds | 48 (36-60) | 29 (17-42) | 39 (29-48) |
| **Gastric tube placement<sup>d</sup>** | 20 (11-29) | 5.7 (0.91-10) | 14 (8.1-19) |
| **Additional feeds by**<br>**caregiver<sup>d</sup>** | 6.9 (1.2-13) | 18 (10-26) | 12 (7.3-17) |

<sup>a</sup>Outcomes are reported as average predicted probabilities (95% confidence interval) for n=141 bCPAP-days, based on a multinomial logistic regression model with robust standard errors.
<sup>b</sup>AKUH: Aga Khan University Hospital
<sup>c</sup>ASH: Abbasi Shaheed Hospital
<sup>d</sup>There were 4 missing values for undisrupted bCPAP therapy, 1 for monitoring, 3 for avoidance of oral feeds, 2 for diet order, 1 for gastric tube placement, and 3 for additional feeds by caregiver.

**Fig 3.**
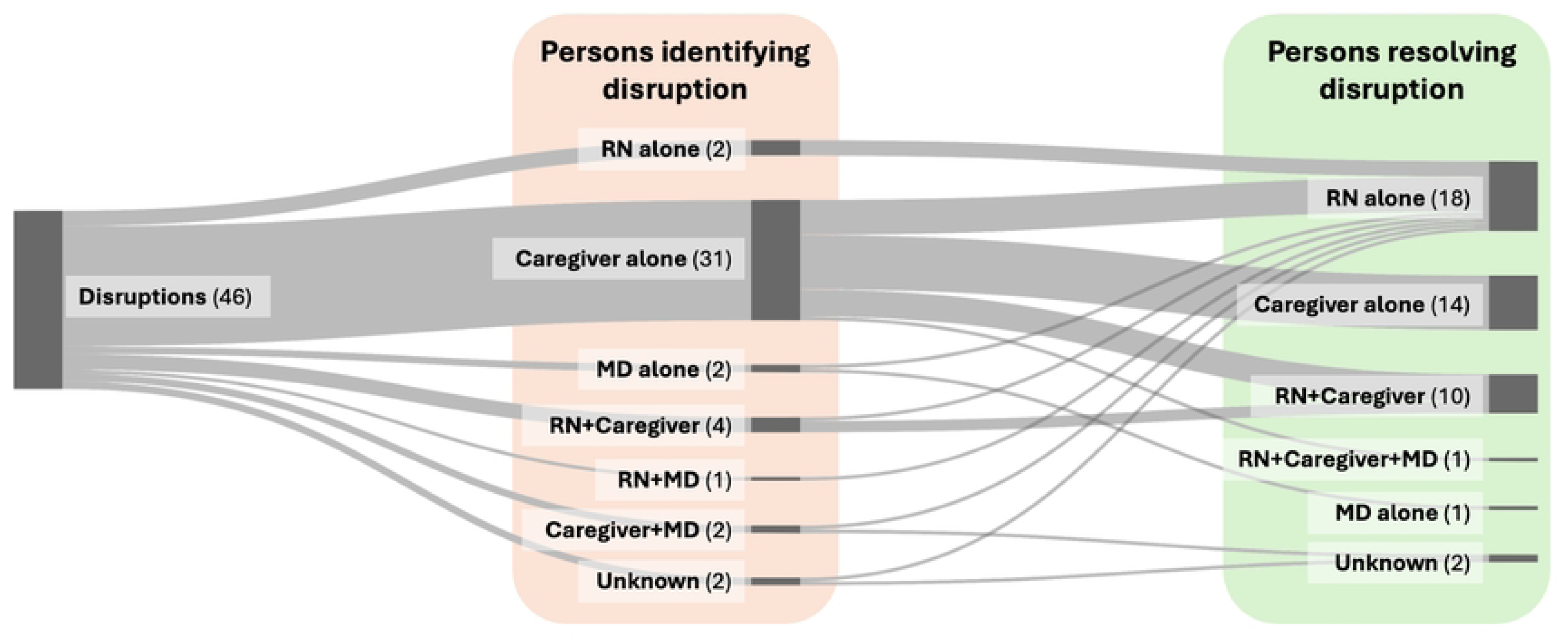
Persons identifying and resolving unplanned disruptions in bCPAP therapy. Frequencies shown in parentheses for each person or group. RN, nurse; Caregiver, familial caregiver at bedside; MD, medical doctor.

Children receiving bCPAP had an average predicted probability of being continuously monitored of 93% (95% CI 88-99%) at AKUH and 12% (95% CI 4.7-20%) at ASH. The overall average predicted probability of a child receiving bCPAP being in a highly monitored area was 85% (95% CI 78-92%) and similar across both sites. The average predicted probability of avoidance of oral feeding among children receiving bCPAP was 46% (95% CI 36-55%) and similar across both sites. Children receiving bCPAP at AKUH had a higher average predicted probability than those at ASH of having a gastric tube placed (20%, 95% CI 11-29% versus 5.7%, 95% CI 0.91-10%) and a lower average predicted probability of caregivers giving additional feeds despite doctors’ orders (6.9%, 95% CI 1.2-13% versus 18%, 95% CI 10-26%) (**Table 2**).

### bCPAP safety and effectiveness

Of 88 children who received bCPAP: 80 (90%) children were followed up through discharge, transfer to another hospital, or death; 9 (10%) experienced an aspiration event and 1 (1.1%) experienced a pneumothorax. 19 (22%) children experienced treatment failure, including 14 (16%) children who left against medical advice and 6 (6.8%) who were escalated to a higher level of respiratory support. Only 1 (1.1%) child died during the follow-up period (**Table 3**). Final disposition and mortality for all enrolled children, including those who did not receive bCPAP therapy, is available in the S4 Table.

**Table 3.** Patient-level safety and effectiveness outcomes, by study site.

| **Outcome, n (%)** | **AKUH (n = 45)** | **ASH (n = 43)** | **Overall (n = 88)** |
| --- | --- | --- | --- |
| **Aspiration** | 6 (13) | 3 (7.0) | 9 (10) |
| **Pneumothorax** | 1 (2.2) | 0 (0.0) | 1 (1.1) |
| **Treatment failure** | 8 (18) | 11 (26) | 19 (22) |
| Death | 1 (2.2) | 0 (0.0) | 1 (1.1) |
| Change to higher respiratory support | 6 (13) | 0 (0.0) | 6 (6.8) |
| Left against medical advice | 3 (6.7) | 11 (26) | 14 (16) |
| **Lost to follow up** | 0 (0.0) | 8 (19) | 8 (9.1) |

## Discussion

In this prospective cohort study of 165 children with severe pneumonia across two hospitals in Pakistan, we identified several gaps in bCPAP implementation fidelity and safety. bCPAP reach varied across our two sites. Among children receiving bCPAP therapy, unplanned disruptions to bCPAP therapy were frequent, some children were cared for in ward settings without resources for continuous monitoring, gastric tubes for stomach decompression were rarely placed, and some children were orally fed—sometimes by doctor’s orders, and sometimes by caregivers despite doctor’s orders. Almost one-fourth experienced bCPAP treatment failure, often due to leaving against medical advice while receiving bCPAP therapy. Our findings highlight opportunities to improve the fidelity and safety of bCPAP therapy in such settings.

bCPAP reach was much higher at ASH, a public hospital with limited access to other non-invasive respiratory support modalities, compared to AKUH, a private hospital where many children were started on high-flow nasal cannula (HFNC) oxygen as an alternative to bCPAP. HFNC is costlier than bCPAP (22), and a 2022 systematic review of five trials comparing HFNC to bCPAP for children under 2 years old with acute bronchiolitis, most of whom would have met eligibility criteria for our study, found a higher risk of treatment failure and longer length of stay in the HFNC group (23). While one recent trial in India demonstrated a lower risk of treatment failure with HFNC compared to bCPAP, this trial used a collapsible bag as its bCPAP water reservoir, which may produce inadequate pressures (24). Considering this evidence, future trials should include implementation outcome measures to better understand which therapies are most effective in which contexts, especially considering that HFNC is significantly costlier than bCPAP.

Oral feeding while on bCPAP was common at both AKUH and ASH, with gastric tube placement rarely performed at both sites. While a 2021 systematic review found that evidence is insufficient to conclude whether oral feeding while on bCPAP is safe (25), others have asserted that the risk of oropharyngeal aspiration is higher in children receiving bCPAP therapy, and oral feeding in the one bCPAP trial may have caused higher deaths in the bCPAP group compared to the control group (11,26). Additionally, our finding that caregivers sometimes gave their children additional feeds despite doctors’ orders is aligned with the findings of other qualitative studies in Malawi that found caregivers’ desire to feed their children orally and negative perceptions of nasogastric feeding (27,28). More studies are needed to confirm the safety of or harm caused by oral feeding while receiving bCPAP and develop strategies to collaborate with caregivers to safely feed their children receiving bCPAP.

While most children receiving bCPAP were in an emergency department, high-dependency unit, or intensive care unit, which benefit from higher staff-to-patient ratios, some children receiving bCPAP at both sites were still in ward settings. Many children, especially at ASH, were not continuously monitored, although the average predicted probability of an unplanned disruption—only one type of bCPAP-related event needing urgent intervention—was over one-third across both sites. While pulse oximetry is available at 79-92% of secondary and 83-96% tertiary care hospitals in low-income and middle-income countries (29), patient-level access to continuous pulse oximetry is almost certainly much rarer. This poses a unique problem: how will children receiving bCPAP, a continuous therapy with potentially high rates of unplanned disruptions or complications requiring urgent intervention, be monitored? Although continuous pulse oximetry is ideal, in settings where this is not feasible, it may be possible to train those who are continuously at the bedside—often caregivers—to recognize bCPAP-related events needing urgent intervention. Indeed, even though they received no formal training, caregivers in our study identified most unplanned disruptions and often worked independently or with nursing staff to resolve the disruption. Future research should explore strategies to educate and involve caregivers in bCPAP-related care to improve safety.

The high frequency of children leaving against medical advice while receiving bCPAP therapy offers another opportunity to improve care by collaborating with caregivers. While we did not systematically analyze reasons for leaving against medical advice in our study, anecdotally reported reasons included dissatisfaction with care, desire to seek care at a different hospital, not wanting a hospital admission, and financial constraints (at the private hospital). Future studies should examine reasons why children receiving bCPAP leave against medical advice, since this poses a risk that is potentially modifiable by improving communication between clinicians and caregivers.

## Strengths and limitations

Our study had several strengths. We based our study on a bCPAP implementation framework derived from our prior realist review (14), focusing on fidelity outcome measures for bCPAP therapy. This allowed us to gain insights that have been sparsely documented in prior bCPAP literature. We enhanced the generalizability of our findings by including two sites with contrasting resource constraints. Our pragmatic use of the WHO clinical criteria for severe pneumonia further enhanced the generalizability of our findings, since chest radiography is not available at many sites that use bCPAP.

It is also important to interpret our study in the context of the following limitations. First, our sampling frame at each site may have introduced selection bias, both because we did not include children who presented outside of our enrollment hours at ASH and because we only enrolled children during four months of the year. We were therefore unable to capture diurnal or seasonal variations in our outcomes. Second, at ASH, due to local clinical protocols, almost all children in respiratory distress were immediately started on bCPAP, without recording vital signs before bCPAP initiation. We were therefore unable to estimate the pre-bCPAP prevalence of hypoxemia and other clinical characteristics of children at ASH. Third, we collected data daily, and some of our outcomes, including disruptions in bCPAP therapy and feeding practices, depended on caregiver and nursing report. Thus, we expect that we may have underestimated the true prevalence of some outcomes, including disruptions and feeding against doctors’ orders. Fourth, while our sites faced resource constraints, even our public hospital site was relatively well-resourced compared to many other settings implementing bCPAP therapy. Future research should confirm our findings in more resource-constrained settings. Our eligibility criteria relied on the WHO definition of severe pneumonia, which is nonspecific, and many children in our study had chest radiographs that were inconsistent with pneumonia. However, chest radiography is not available at many resource-constrained hospitals; using WHO criteria may increase the generalizability of our findings.

## Conclusion

We identified several gaps in bCPAP reach and implementation fidelity, including a lack of continuous patient monitoring, a high frequency of unplanned disruptions in bCPAP therapy, and potentially risky oral feeding practices. These may be modifiable by individual- and team-targeted strategies to reduce bCPAP-related complications and pneumonia-related child deaths.

## Data Availability

The datasets used in our study contain potentially identifying and sensitive patient information, and we have not received approval from the adjudicating ethics committees to publicly share them. Individuals requesting dataset access may contact Dr. Qalab Abbas or Dr. Fyezah Jehan.

## Acknowledgements

We thank Dr. Veronika Shabanova, Associate Professor of Pediatrics and of Biostatistics, Yale School of Medicine, for her help in guiding our statistical analysis.

## Supporting Information Captions

**S1 Appendix.** Strengthening the Reporting of Observational Studies in Epidemiology (STROBE) checklist.

**S2 Appendix.** REDCap data collection forms.

**S3 Table.** Child and caregiver characteristics for children who did and did not receive bCPAP, by study site.

**S4 Table.** Final disposition and mortality for children who did and did not receive bCPAP, by study site.

